# Dietary patterns and nutritional composition of packed lunches in early years education settings

**DOI:** 10.64898/2026.03.03.26347536

**Authors:** Sanjoy Deb, Mitzi Harris, Michelle Hawkins, Michelle Wisbey, Stacy Randall, Kay Aaronricks

## Abstract

**Background:** Packed lunches are a common feature of early childhood food provision, yet evidence describing their nutritional composition in early years settings remains limited. Understanding the foods provided during this developmental period is important, given the potential influence of early dietary exposures on later health.

**Aim:** To characterise the composition, nutritional quality, cost, and dietary patterns of packed lunches brought from home in Early Childhood Education and Care settings, and to examine variation by child age and area-level deprivation.

**Methods:** A cross-sectional analysis was conducted using a remote food photography method to assess packed lunches provided for children aged 1–4 years attending early years settings across Essex, UK. Food items were categorised into predefined groups, and nutrient composition was estimated. Area-level deprivation was determined using the English Index of Multiple Deprivation (2019). Non-parametric tests assessed between-group differences. Principal components analysis (PCA) was used to identify patterns of co-occurring foods.

**Results:** A total of 389 packed lunches were analysed. Starchy foods (82%), fruit (81%), dairy or alternatives (72%), and savoury snacks (74%) were commonly provided, while vegetables were less frequent and fish was rarely observed (1.5%). Overall, 97.7% of lunches contained at least one ultra-processed food (UPF), with a median of three UPF items per lunch and 74% of total energy derived from UPFs. Median energy provision was 400 kcal (IQR 309–518). Nutrient composition was broadly similar across deprivation groups, although cake and biscuit counts and UPF item counts were modestly higher in more deprived areas. The median estimated lunch cost was £1.79 and did not differ by deprivation.

**Conclusions:** Packed lunches in early years settings frequently contained ultra-processed foods and showed considerable variability in nutritional quality. Socioeconomic differences were limited, suggesting that contemporary packed lunch practices may reflect influences operating across population groups. Further research across diverse regions is warranted to better understand the provision of packed lunches and their implications for early dietary exposure.

## Introduction

Early Childhood Education and Care (ECEC) settings are increasingly recognised as a critical environment for shaping children’s health and reducing social inequalities ^(1)^. In England, uptake of early education is high among those eligible for government-funded provision, with approximately 95% of 3–4-year-olds attending ECEC, where children spend an average of around 22 hours per week. Recent policy changes have further increased the relevance of ECEC, with the expansion of funded childcare to include children aged 9 months to 2 years expected to increase both attendance and time spent in early years provision at younger ages. As a result, many young children consume a substantial proportion of their daily food intake within nurseries, preschools, and childminders’ settings ^(2)^. Children attending a typical full-day EYS may eat two to three meals and two snacks, accounting for up to 90% of daily energy intake, making EYS a particularly important context for health promotion and dietary intervention ^(2)^.

Adequate nutrition during early childhood is essential for healthy growth, development, and disease prevention ^(1,3)^. Dietary quality among young children in high-income countries remains suboptimal, with many failing to consume adequate fruits and vegetables while frequently consuming sugar-sweetened drinks ^(4)^. In England and Scotland, approximately 22% of children are classified as overweight or obese by the time they reach reception age ^(5)^. These challenges are compounded by socioeconomic inequalities, with nearly 19% of UK households with children experiencing food insecurity, limiting access to affordable, balanced diets and disproportionately affecting families living on lower incomes ^(6)^. Given both the number of eating occasions in ECEC and the increasing proportion of children attending from infancy, it is important to understand how the nutritional environment within ECEC influences children’s diets and health outcomes.

Unlike primary schools, which are required to comply with national school food standards, early years providers in England are not subject to mandatory nutritional regulations for meals. Food provision practices vary widely, with some ECEC’s preparing meals and snacks on-site, while others provide no food at all and require families to send packed lunches for their children. Although voluntary guidance, including the Eat Better, Start Better guidelines ^(7)^ and Department for Education Early Years Foundation Stage (EYFS) Nutrition Guidance, is available to support healthy food provision ^(8)^, uptake is not compulsory. This reliance on voluntary guidance has contributed to variability in food provision and uneven access to nutrition support across settings ^(9)^. Updated statutory guidance further clarifies that families must be able to access funded early years education without additional food-related charges, allowing parents to provide packed lunches in place of meals offered by the setting (Department for Education, 2025). This policy context suggests that packed lunches may become an increasingly common source of dietary intake in early years settings, reinforcing the need to examine their nutritional composition.

Evidence from school-aged children shows that packed lunches are often nutritionally inferior to school-prepared meals, with lunchboxes frequently containing energy-dense, nutrient-poor items such as sweet snacks, confectionery, and salty foods, alongside limited provision of fruit and vegetables ^(10)^. In contrast, research examining packed lunches among children under five remains comparatively scarce. A recent study of lunchbox provision in early years settings in Sheffield provides important insight across multiple dimensions, showing that parent-packed lunches commonly exceeded recommended levels for energy, free sugars, sodium, and several macronutrients, including total fat and saturated fat ^(11)^. Clear socioeconomic differences were also evident, with packed lunches provided in more deprived areas containing fewer fruits and vegetables and a greater emphasis on items such as meat products, savoury snacks, cakes and biscuits, chocolate or sugar confectionery, and squash or fruit-based drinks ^(11)^. The same study highlighted the economic dimension of lunchbox provision, indicating that lunchboxes of higher nutritional quality were, on average, less costly than those of lower nutritional quality, whereas lunches dominated by packaged snack products tended to be more expensive overall. These findings challenge the assumption that healthier packed lunches necessarily incur greater costs and suggest that food choice, particularly reliance on packaged snack items, may be a key driver of both nutritional quality and affordability. This pattern aligns with broader concerns about the increasing contribution of ultra-processed foods to children’s diets ^(12)^. Indeed, Pearce and Wall ^(11)^ reported that ultra-processed foods accounted for 65.5% of the total energy provided in 185 early years lunchboxes.

Despite growing recognition of early childcare as a key setting for nutritional intervention, recent evidence remains limited to a few geographical areas in the UK. Previous studies of young children’s lunches have primarily focused on centre-based providers (nurseries and preschools), whereas home-based childminders, who also care for a portion of the under-5 population, have been underrepresented in research. Therefore, this project aims to characterise the composition, nutritional quality, cost, and underlying patterns of packed lunches brought from home in early years settings throughout Essex, and to examine how these patterns vary by setting, child age, and socioeconomic context.

## Methods

### Study Design and Setting

This cross-sectional observational study examined packed lunches brought from home for children aged 1–4 years attending early years settings across Essex, UK. Data were collected between November 2024 and January 2025, capturing lunch provision on a typical day in the setting. Participating settings included nurseries, preschools, and childminders, representing a geographically diverse sample across the county. Essex is characterised by socioeconomic diversity spanning urban, rural, and coastal communities, providing an opportunity to capture packed lunch provision across varied local contexts. No guidance was provided to caregivers or settings regarding lunchbox contents prior to or during the study. Ethical approval was obtained from the institutional research ethics committee at Anglia Ruskin University (Reference: ETH2425-0326), and written informed consent was obtained from parents/carers and participating settings.

### Early Years Setting Recruitment

Early years settings across Essex were invited to participate via an email distributed by the local authority to all registered providers (approximately N = 1344). Interested settings were invited to attend either an in-person or virtual workshop held during and outside of normal working hours, during which study procedures were discussed to ensure feasibility within routine practice and guidance on data collection was provided. Settings were eligible to participate if they cared for children aged 1–5 years and permitted packed lunches brought from home. Within participating settings, all children who brought a packed lunch on the day of data collection were eligible for inclusion. Lunchboxes were assessed on a single occasion per child to capture food provision under usual conditions.

### Collection of photos

Lunchbox images were captured by educators within participating settings on a typical day selected by the setting. Data collection procedures were introduced during the study workshop, and settings were provided with a standardised instruction document to support implementation. Educators were asked to place lunchboxes or individual food items on an A4 sheet of paper to provide a reference for estimating portion size. Images were taken before any food was consumed, with all items removed from containers and arranged so that contents were clearly visible wherever possible. A single photograph capturing all food items was recorded for each lunchbox. The use of photographic methods to assess the provision of packed lunches has previously been validated in preschool populations ^(13)^.

### Pack lunch composition

Analyses were conducted at the lunchbox level using photographic assessment data collected from participating early years settings. Foods were categorised into twelve pre-specified food groups: fruit (fresh, dried, or tinned), vegetables, starchy foods, non-dairy protein sources, dairy or alternatives, meat products, processed cereal bars or processed fruit items, savoury snacks, cakes or biscuits, chocolate or sugary confectionery, squash or fruit juice, and fish. Food-group counts reflected the number of distinct items observed in each lunchbox, with zero indicating the absence of items. Food coding was conducted by two trained nutrition researchers.

### Nutrient coding and analysis

Nutrient analysis was conducted at the lunchbox level using Nutritics nutritional analysis software (Nutritics Ltd., Dublin, Ireland). Portion sizes were estimated independently from the images by two trained nutrition researchers, with discrepancies resolved through discussion to reach consensus. Where composite or mixed foods were clearly identifiable (e.g., sandwiches), items were disaggregated into their constituent ingredients prior to analysis. Lunchboxes containing items that could not be reliably identified from the images were excluded from nutrient analysis. For each lunchbox, total energy, macronutrients, and fibre were calculated by summing values across all identified food items.

### Lunchbox Healthiness Score

A lunchbox healthiness score (range 0–11) was calculated to reflect alignment with food-based recommendations. The scoring approach was informed by previous research applying the Eat Better, Start Better guidelines ^(11)^. One point was assigned for the presence of each recommended food group (starchy foods, fruit, vegetables, non-dairy protein, and dairy or alternatives), and one point for the absence of food groups recommended to limit (meat products, processed fruit items or cereal bars, savoury snacks, cakes or biscuits, chocolate or sugary confectionery, and squash or cordial). Higher scores indicated greater alignment with recommendations. Lunchboxes scoring ≥8 were classified as healthier, while those scoring ≤7 were classified as less healthy.

### Ultra-Processed Food Classification

Food items were classified according to the NOVA food classification system, which categorises foods into four groups based on the extent and purpose of industrial processing ^(14)^. For the purposes of this study, analyses focused on foods classified as Group 4 (ultra-processed foods; UPFs). Coding was conducted using detailed information recorded from the images, including food type, product description, and brand where identifiable. Items not explicitly described within the NOVA framework were classified with reference to Open Food Facts and prior research applying NOVA in UK populations (^(15)^; [Open Food Facts, 2025](https://world.openfoodfacts.org/)). UPF coding was completed by one researcher and checked by a second trained nutrition researcher, with queries resolved through discussion to reach consensus.

### Cost Analysis

Estimated food costs were derived using retail price data collected in July 2024. Unit prices were obtained primarily from Tesco’s online grocery platform to reflect costs within a widely used UK supermarket. For staple unbranded items (e.g., fresh fruit and vegetables, bread, ham, and cheese), the lowest-priced own-brand products were selected. Where branded items were identifiable from images, the corresponding product price was applied. Promotional prices were included where applicable to reflect typical consumer purchasing conditions. Total lunchbox cost was calculated by summing the estimated cost of all identified items.

### Area Deprivation

Area-level deprivation was determined using the Income Deprivation Affecting Children Index (IDACI) 2019, a domain of the English Indices of Multiple Deprivation that reflects the proportion of children aged 0–15 years living in income-deprived households within a given area. Postcodes for participating early years settings were linked to lower-layer super output areas (LSOAs), from which national IDACI deciles were assigned. Settings were categorised into higher-deprivation (deciles 1–5) and lower-deprivation (deciles 6–10) groups to enable comparison across socioeconomic context.

### Statistical Analysis

Analyses were conducted using IBM SPSS Statistics (Version XX; IBM Corp., Armonk, NY, USA). Records were included where food items could be identified and the relevant stratification variable was available. Continuous variables were not normally distributed and were summarised using medians and interquartile ranges (IQRs), alongside means and standard deviations (SDs) where appropriate. Food group prevalence is presented as the percentage of lunchboxes in which each item was observed (count >0). Between-group differences in continuous outcomes were assessed using non-parametric tests. Wilcoxon rank-sum tests were used to compare differences between age groups and area-level deprivation groups (IDACI), while Kruskal–Wallis tests were applied for comparisons involving more than two groups. Differences in categorical variables were examined using chi-squared tests, with Holm-adjusted post hoc comparisons conducted where overall associations were identified. All tests were two-sided, with statistical significance set at p < 0.05.

Dietary patterns were explored using principal component analysis (PCA) based on eleven pre-specified food-group variables: fruit, vegetables, starchy foods, non-dairy protein sources, dairy or alternatives, meat products, processed cereal bars or processed fruit items, savoury snacks, cakes or biscuits, chocolate or sugary confectionery, and squash or fruit juice. Fish was excluded due to low prevalence. PCA was performed on the correlation matrix using lunchboxes from children aged 1–4 years. Missing values were imputed using variable means. Sampling adequacy was assessed using the Kaiser–Meyer–Olkin statistic and Bartlett’s test of sphericity. Components were retained based on eigenvalues >1 and interpretability, and varimax orthogonal rotation was applied to aid interpretation. Rotated component scores were calculated for each lunchbox.

## Results

### Early Years Settings Characteristics

A total of 55 ECEC settings participated in the study, including 26 childminders, 8 day nurseries, 21 preschools. Overall, 411 lunchbox photographs were received; however, 22 were excluded because food items could not be clearly distinguished, leaving 389 packed lunches for children aged 1–4 years for inclusion in the analysis. Age data were available for 346 lunches: 112 (28.8%) were for children aged 1–2 years, 234 (60.2%) for those aged 3–4 years, and 43 (11.1%) were missing. Based on area-level deprivation (IDACI), data were available for 388 lunches, with 194 (49.9%) from settings located in more deprived areas (deciles 1–5) and 194 (49.9%) from less deprived areas (deciles 6–10); deprivation data were missing for one lunch (0.3%).

### Lunchbox Composition

Across all lunches (n = 389), the most commonly observed food groups were starchy foods (82.3%), fruit (81.2%), savoury snacks (74.0%), and dairy or alternatives (72.2%). Fish was rarely observed (1.5%). Lunch composition was broadly similar across deprivation groupings and age groups. A between-group difference was observed only for cakes and biscuits, which were more commonly present in lunches from more deprived areas (41.8%) compared with less deprived areas (27.8%) (p = 0.004). No other food groups differed at the p < 0.05 level (Table 1).

**Table 1.** Composition of packed lunches in early years settings: including overall mean item count and percentage presence overall, by child age, and by area-level deprivation.

| Item | Overall: Any present (%) | Overall: Mean ± SD | Age: 1-2 | Age: 3-4 | IDACI: 1-5 | IDACI: 6-10 |
| --- | --- | --- | --- | --- | --- | --- |
| Starchy food | 82.26 | 1.14 ± 0.77 | 82.14% | 83.76% | 79.38% | 85.05% |
| Fruit | 81.23 | 1.31 ± 0.97 | 82.14% | 78.63% | 83.51% | 79.38% |
| Savory snacks | 74.04 | 0.99 ± 0.80 | 75.9% | 73.50% | 72.68% | 75.77% |
| Dairy or alternatives | 72.24 | 1.04 ± 0.83 | 72.32% | 73.08% | 75.26% | 69.07% |
| Meat products | 44.47 | 0.55 ± 0.70 | 45.54% | 45.30% | 44.33% | 44.3% |
| Chocolate/sugary confectionery | 42.93 | 0.52 ± 0.68 | 37.50% | 46.58% | 44.85% | 40.72% |
| Non-dairy protein | 38.05 | 0.46 ± 0.64 | 39.29% | 38.89% | 39.69% | 36.08% |
| Cake/biscuit | 34.70 | 0.38 ± 0.57 | 33.04% | 35.47% | 41.75% | 27.84% |
| Vegetables | 28.28 | 0.37 ± 0.67 | 25.00% | 28.63% | 24.74% | 31.96% |
| Processed cereal bars/fruit | 16.71 | 0.21 ± 0.53 | 23.21% | 13.68% | 13.92% | 19.59% |
| Squash/fruit juice | 7.46 | 0.08 ± 0.28 | 7.14% | 6.84% | 7.73% | 7.22% |

## Nutrient Composition of Lunchboxes

Across all lunches, median energy provision was 400 kcal (IQR 309–518). Median carbohydrate provision was 53.6 g (40.6–69.9), total sugars 22.5 g (14.1–32.6), fibre 3.6 g (2.5–5.4), protein 12.3 g (8.2–17.1), and fat 13.4 g (9.2–21.0). Nutrient composition did not differ by area-level deprivation, with no statistically significant differences observed for energy or any nutrient outcome (all p > 0.05).

Some variation was observed by age group. Lunches provided for children aged 1–2 years contained higher fibre (median 4.1 vs 3.4 g; p < 0.001) and total sugars (median 23.8 vs 21.4 g; p = 0.013), but lower fat content (median 11.9 vs 14.8 g; p = 0.013) compared with lunches for children aged 3–4 years. No age-group differences were observed for energy, carbohydrate, or protein provision.

### Ultra-processed foods

Overall, 380 of 389 lunches (97.7%) contained at least one ultra-processed food (UPF) item. The mean UPF count was 3.08 ± 1.46, with a median of 3 items (IQR 2–4). UPF counts were higher among children aged 3–4 years than those aged 1–2 years (mean 3.25 vs 2.79; p = 0.009). Counts also differed by deprivation grouping (p = 0.017), with modestly higher counts observed in the more deprived group (mean 3.25 ± 1.59) compared with the less deprived group (mean 2.92 ± 1.31). The proportion of total lunch energy derived from UPFs was high, with a median of 74.2% (58.9–88.6). UPF energy contribution did not differ by deprivation grouping (p = 0.716). A small difference was observed by age group, with lunches for children aged 3– 4 years showing a slightly higher median UPF energy percentage than those for children aged 1–2 years (75.0% vs 71.6%; p = 0.046).

### Estimated lunch cost

Estimated cost data were available for 389 lunches, with a median cost of £1.79 (£1.28–£2.41). Costs did not differ by deprivation grouping (p = 0.547). Lunches for children aged 1–2 years had a slightly higher median estimated cost than those for children aged 3–4 years (£1.93 vs £1.69; p = 0.018).

### Overall lunchbox healthfulness score and dietary patterns

Median lunch score was 7 (IQR 6–8), indicating moderate alignment with food-based recommendations. Approximately one-third of lunches (33.7%) met the threshold for classification as healthier (≥8 recommendations). Scores did not differ by age group (p = 0.951) or deprivation grouping (p = 0.646).

The overall Kaiser–Meyer–Olkin (KMO) measure of sampling adequacy was 0.55, indicating relatively weak common variance across the variable set. In contrast, Bartlett’s test of sphericity was statistically significant (p < 0.001), indicating that the correlation matrix was not an identity matrix and that sufficient intercorrelation existed to justify exploratory dimensional reduction. Therefore, while the data were suitable for PCA, the resulting components were expected to be modestly defined and should be interpreted cautiously.

Principal component analysis identified six components with eigenvalues greater than 1, which together accounted for 74% of the variance in food-group provision. The first component explained 19.1% of the variance, followed by the second (15.4%), third (10.8%), fourth (10.1%), fifth (9.5%), and sixth (9.1%). These components were interpreted as patterns of co-occurring food groups within lunchboxes and labelled accordingly: (1) protein-rich foods and savoury snacks, (2) sweet snacks and confectionery, (3) sweet drinks and processed snack items, (4) starchy foods with vegetables, (5) fruit and sweet drinks contrasted with savoury snacks, and (6) dairy and accompanying snack items.

The first pattern was characterised by strong positive loadings for non-dairy protein (0.95) and meat products (0.95), with a smaller contribution from savoury snacks (0.34). The second pattern showed high positive loadings for cakes and biscuits (0.86) and chocolate or sugary confectionery (0.86), alongside a modest negative loading for vegetables (−0.31). The third pattern was defined by processed cereal bars or processed fruit items (0.83) and squash or fruit juice (0.63), indicating the co-occurrence of packaged snack items and sweet drinks. The fourth pattern was characterised by strong positive loadings for starchy foods (0.79) and vegetables (0.69). The fifth pattern showed a strong positive loading for fruit (0.82) and a moderate positive loading for squash or fruit juice (0.42), together with a negative loading for savoury snacks (−0.41). The sixth pattern was dominated by dairy or alternatives (0.92), with a smaller positive contribution from savoury snacks (0.42).

**Figure 1.**
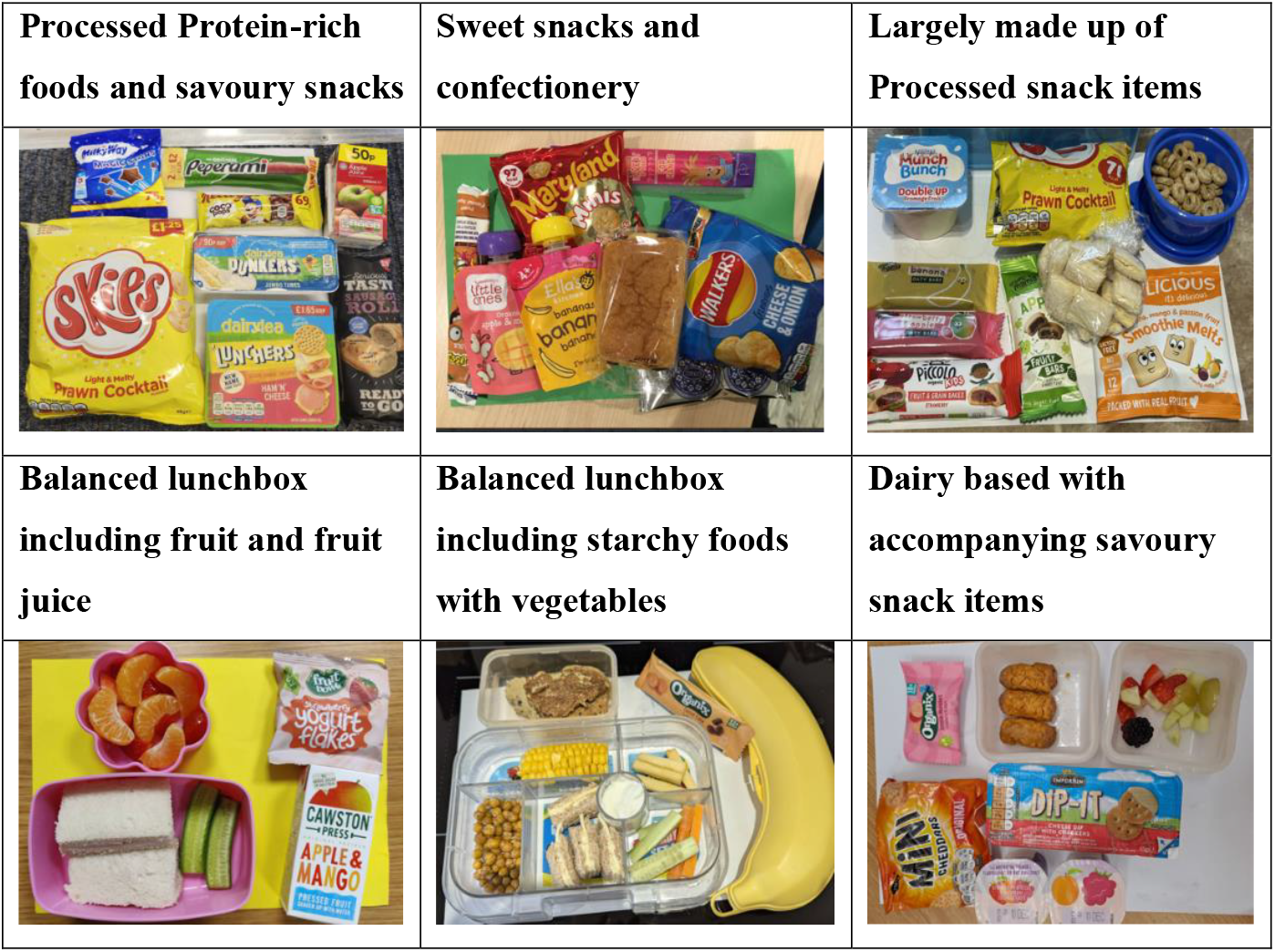
Example packed lunches illustrating dietary patterns identified through principal component analysis (PCA).

## Discussion

This study characterises packed lunches brought from home in early years settings, extending the limited UK evidence base beyond school-aged populations. Using photographic assessment across a county-wide sample, we found that lunches commonly included starchy foods, fruit, dairy items, and savoury snacks, but were also marked by the near-ubiquitous presence of ultra-processed foods (UPFs). While overall composition and nutrient profiles were broadly similar across socioeconomic groups, differences emerged by child age, particularly in relation to snack foods, fibre, sugars, fat, and the contribution of UPFs to energy and cost. The identification of distinct dietary patterns further suggests that packed lunches are not homogeneous but reflect recurring combinations of foods. These findings highlight packed lunches as an important yet under-examined component of early childhood diets, with implications for dietary quality and exposure to UPFs during a critical period of development. These findings are particularly important in a sector described as under-resourced and insufficiently prioritised, despite its central role in shaping young children’s dietary exposures ^(9)^.

The median energy provision observed in the present study (400 kcal) closely aligned with the lunch energy target outlined in the Eat Better, Start Better guidance (369 kcal) ^(7)^, but was lower than estimates reported in packed lunch research conducted in early years settings in Sheffield (524 kcal) and somewhat below values from a national study spanning fifteen local authority or health board regions (426–471 kcal) that included both setting-provided meals and parent-provided lunches ^(11,16)^. This difference may partly reflect dietary assessment methods. While the Sheffield study employed weighed measures of food intake, the remote food photography approach used here captures foods in naturalistic settings but may produce more conservative estimates where portion sizes are uncertain. Importantly, our findings are comparable to those from the national study that applied a similar photographic method ^(13)^, supporting the credibility of these estimates. It is also notable that the national analysis combined setting meals with lunchboxes, despite evidence that parent-provided lunches often contain larger portions than foods prepared within early years settings ^(16)^. Meal size is an important consideration in early childhood, as longitudinal evidence indicates that each additional 10 kcal consumed per meal is associated with a 7% faster rate of weight gain among children aged 2–5 years ^(17)^. Although this increase appears small, repeated exposure across multiple eating occasions may contribute meaningfully to cumulative energy intake, highlighting the importance of understanding the foods and dietary patterns underpinning energy provision within packed lunches.

The near-universal presence of UPFs within packed lunches is consistent with growing evidence that such foods dominate children’s diets from an early age in the UK. Analyses of national dietary data show that UPFs account for more than half of total energy intake among UK children, with this proportion increasing across early and mid-childhood ^(12,18,19)^. In school-aged populations, packed lunches contain a higher proportion of energy from UPFs than school-provided meals, driven largely by processed breads, drinks, and sweet and savoury snacks ^(18)^. Evidence from younger age groups suggests similar patterns; packed lunches in early years settings in Sheffield derived 65.5% of total calories from UPFs ^(11)^, while a large cohort study reported that children aged 21 months consumed 46.9% of daily energy from UPFs ^(12)^. Although foods classified as ultra-processed using the NOVA framework are often associated with poorer health outcomes, interpretation requires nuance. Many UPFs are hyperpalatable, energy-dense, and high in sugar or sodium, characteristics linked to obesity and long-term cardiometabolic risk ^(20,21)^. However, excluding all UPFs may reduce intake of key nutrients obtained through fortified foods and overlook gains achieved through fortification policy ^(22)^. For example, high-fibre cereals were identified as major contributors to UPF intake among toddlers ^(12)^. Nonetheless, the present results reinforce concerns that packed lunches may represent an important source of UPF exposure during a formative stage of dietary development.

An examination of lunchbox composition provides important context for interpreting the nutritional profile of packed lunches in early years settings. Consistent with previous UK research, fruit and dairy items were commonly included, whereas vegetables were less frequently observed. In a weighed study of packed lunches in Sheffield, fruit was present in 76.2% of lunches and vegetables in 38.9%, with dairy or alternatives included in 80.5% ^(11)^, a distribution broadly comparable to that observed here. Snack-type items were also frequently present alongside core food groups, and individually pre-packaged foods accounted for a substantial proportion of items. Assessment of overall dietary quality indicated considerable variability in how well lunches aligned with food-based recommendations, with approximately one-third meeting the threshold for classification as healthier. Evidence from UK primary school populations similarly indicates that the quality of packed lunches has remained variable over time, with relatively few meeting the established nutritional standards despite modest improvements ^(23)^.

Principal components analysis further supported these observations by showing that foods commonly appeared together within lunchboxes, suggesting that lunches may follow recognisable patterns of provision. Although this pattern-based analysis is commonly used in nutrition research to examine daily dietary patterns^(24)^, research on lunchbox patterns remains limited and lacks comparisons. Social practice theory offers a useful lens for explaining the emergence of food patterns, conceptualising lunchbox preparation as part of everyday food routines shaped by activities such as planning, shopping, and packing ^(25)^. Within these routines, foods may be grouped through habit rather than intentionally paired. Evidence also indicates that while many parents consider nutritional guidance when preparing lunchboxes, they often balance this with practical factors including children’s preferences, cost, convenience, and time ^(26)^. Parental food practices influence children’s eating behaviours ^(27)^, yet nutrition knowledge alone does not necessarily translate into healthier food provision ^(28)^. Collectively, these studies suggest that the combinations observed across lunchboxes are shaped by multiple interacting influences within the wider food environment. Although the PCA findings should be interpreted cautiously, given modest shared variance across food groups, recurring food combinations indicate that lunchboxes may reflect established provisioning habits rather than being assembled solely in response to dietary guidance.

Differences by socioeconomic context were less pronounced than reported in some earlier UK early years research. Lunchbox composition was broadly similar across deprivation groupings in the present study, with a between-group difference observed only for cake and biscuit items, which were more commonly present in lunchboxes from more deprived areas (41.75% vs 27.84%). A similar pattern was observed for ultra-processed foods, with slightly higher item counts in the more deprived group (mean 3.25 ± 1.59) compared with the less deprived group (mean 2.92 ± 1.31), although the overall percentage contribution of UPFs to energy did not differ between groups. In contrast, a weighed food study in Sheffield ECEC found that lunches provided in areas of higher deprivation were less likely to contain fruit and vegetables and more likely to include savoury snacks, confectionery, and sweetened drinks, alongside lower overall healthy eating scores ^(11)^. Feeding practices can reflect class-based expectations surrounding “good” parenting, alongside material constraints that shape everyday food decisions ^(29)^. For example, some caregivers prioritise foods they are confident children will eat in order to avoid waste, particularly where financial resources are limited ^(29)^. More broadly, lunchboxes have been described as reflecting wider food system pressures, including the increasing availability and normalisation of ultra-processed and convenience foods ^(30)^. Taken together, the relatively small socioeconomic differences observed here should not be interpreted as evidence of the absence of structural influence. They may indicate that contemporary packed lunch practices are shaped by constraints that operate across socioeconomic groups. This interpretation aligns with early years stakeholder’s perspectives ^(9)^, suggesting that many challenges related to children’s diets arise from broader systemic conditions rather than from parental knowledge or behaviour alone. Indeed, longitudinal evidence indicates that reductions in investment in early years services are associated with higher rates of childhood obesity ^(31)^, highlighting the role of structural conditions in shaping children’s health trajectories.

This study should be considered in light of several limitations. First, dietary assessment was based on the remote food photography method, which captures foods as provided rather than consumed and may therefore overestimate actual intake if items were not eaten. However, this approach offers a valuable advantage in early years research by enabling observation of real-world food provision without placing additional burden on educators. Photographic methods have also been shown to provide reasonably accurate estimates of portion size when direct weighing is not feasible ^(13)^, supporting their suitability for multi-site, naturalistic studies. Second, although the county-wide sample enhances ecological validity, the findings may not be fully generalisable to all UK early years contexts, particularly given regional variation in food environments and provision practices. Further nationally representative studies are therefore needed to determine whether the patterns identified here reflect broader trends in early childhood diets or are shaped by local factors. Third, the cross-sectional design limits the ability to infer causality or examine how lunchbox provision may change over time. However, the study’s primary aim was descriptive, making this design appropriate for establishing an initial evidence base. Longitudinal research could usefully explore how packed lunch provision evolves across early childhood and whether early exposure to particular dietary patterns persists into later childhood. Finally, area-level deprivation was used as a proxy for socioeconomic status and may not fully capture household-level influences such as income, ^(32)^parental education, or cultural practices. While indices such as the Income Deprivation Affecting Children Index (IDACI) provide a useful contextual indicator of the socioeconomic environment in which children live, they reflect area characteristics rather than the circumstances of individual families. Evidence suggests that area-level indicators are commonly used in early years research but may underestimate associations with child health outcomes compared with individual-level measures^(32)^. Despite this limitation, area-based indices remain widely used in public health research and provide a meaningful indicator of structural conditions shaping food environments and access to resources.

This study provides new evidence on the composition of packed lunches brought from home in early years settings. While many lunches included foods consistent with dietary guidance, they were frequently characterised by high levels of ultra-processed foods and considerable variation in overall nutritional quality. The presence of recurring food combinations suggests that lunches may reflect recognisable patterns of provision rather than isolated food choices. Socioeconomic differences were limited, with modest variation observed in specific discretionary items and UPF exposure rather than in overall lunch composition. Together, these findings indicate that packed lunches form an important part of early childhood food environments and contribute to children’s dietary exposures during a key stage of development. Further research across a wider range of settings and regions is warranted to better understand how packed lunch provision varies and how early dietary patterns relate to later health outcomes.

## Data Availability

All data produced in the present study are available upon reasonable request to the authors

